# Effect of Physical Activity on Olfactory Acuity: A Systematic Review

**DOI:** 10.1101/2022.09.30.22280564

**Authors:** Mathieu Cournoyer, Alexandre-Charles Gauthier, Alice Maldera, Fabien Dal Maso, Marie-Eve Mathieu

## Abstract

Olfactory acuity, which includes detection thresholds, identification and appreciation/intensity, seems to decline with aging, obesity, and various neurological disorders. Knowing that the sense of smell influences energy intake, the interest in protecting this sense is constantly increasing. Physical activity might be a key intervention to counteract the loss of olfactory function. This systematic review aims to explore the literature on the effect of physical activity on olfactory acuity. The search strategy consisted of using index terms and keywords in MEDLINE, EMBASE, EBM Reviews – Cochrane Central Register of Controlled Trials, CINAHL, SPORTDiscus, and Web of Science search engine. Data from 17 studies that include 10 861 participants revealed that physical activity has improved olfactory thresholds, identification, and intensity. More precisely, chronic physical activity seemed to have better effects on olfactory components than acute practice. Even though this review clarified evidence about the effect of physical activity on the sense of smell, better methodological consistency is needed across studies such as standardized experimental conditions, the time of the day data are collected, and similar relative energy intake between participants to produce more robust results.

## 1 Introduction

Olfactory acuity is the ability to discriminate the presence or the absence of an odor. This concept could be divided into three components: odor detection threshold, odor identification, and odor intensity. The odor detection threshold represents the lowest odor concentration that the participant can perceive. Odor identification highlights the participant’s ability to discriminate the type of odor. Finally, odor intensity represents the degree of potency or vigor perceived by a participant to a certain type of odor. Also, odor intensity is usually inversely proportional to odor appreciation since a high odor intensity might feel uncomfortable while breathing. This main concept, with its three components, is often correlated with odor concentration. Indeed, the higher the concentration, the easier it should be to detect and identify the odor, and the higher the intensity.

Unfortunately, olfactory acuity seems to decrease in several situations, such as in aging, Parkinson’s and Alzheimer’s diseases, and people living with obesity [1-9]. This deterioration of the sense of smell can affect diet differently in each person. For example, there may be a decrease in the feeling of hunger, a lower quality energy intake, or even a lower protein consumption which could increase the loss of autonomy [10, 11]. In addition, there are also relations between loss of smell and social isolation [11-13], reduced cognitive functions [11, 14-18], fragile mental health [11, 19], and nutritional problems [10, 11]. Adequate energy intake is crucial to execute daily tasks with full autonomy or to perform a more intense activity, such as sports or other forms of exercise [11, 20]. Also, food consumption is directly connected to environmental stimuli such as the appearance of the food, its smell and the social context [21]. Correspondingly, olfactory sensitivity could affect energy balance by influencing energy intake. A study by Ginieis, Abeywickrema [22] suggested that a decreased olfactory sensitivity would lead to greater snack consumption between meals. Considering that many snacks consumed are energetically dense [23], it is possible to observe the simultaneous increase between the number of obesity cases and the growing habit of consuming snacks daily [24, 25]. Consequently, the sense of smell is an essential component of the quality of life, and that is why it needs to be preserved.

Interestingly, physical activity (PA) seems to protect against the deterioration of the sense of smell. Indeed, treating the loss of olfactive acuity in Parkinson’s patients using PA might offer the same benefit as pharmacologic therapies, and this type of intervention showed to improve not only the olfactive function, but also motor symptoms [26, 27]. It is also possible to look into the physiological process of the olfactory loss to better understand the PA’s impact. A study comparing patients with type 2 diabetes to healthy adults found that high blood pressure could decrease olfactory acuity for both groups [28]. Also, a Schiffman [9] study investigating olfactory loss documented some physiological changes that occurred with aging such as the loss of cells and neurotransmitters in the hippocampus and the olfactory bulb in the brain. These changes could explain olfactory loss and could be positively modified by PA. Erickson, Voss [29] showed that chronic PA reduced the effect of normal aging of hippocampus volume. Knowing that this cerebral structure is involved in the olfactory function, there could be a link between PA and olfactory acuity.

A deeper understanding of the effects of PA on olfactory acuity would be beneficial to highlight the mechanisms modulating energy intake. This systematic review, therefore, aims to highlight the scientific consensus on the influence of PA on olfactory acuity. We will explore the effect of different types of PA on the three component of olfactory acuity, namely, odor detection thresholds, odor identification and odor intensity.

## 2 Methods

### 2.1 Search Strategy

This systematic review followed the “Preferred Reporting Items for Systematic Reviews and Meta-Analysis” (PRISMA) guidelines. Six databases were used to perform the search: Medline (1946-present), Embase (1974-present), EBM Reviews – Cochrane Central Register of Controlled Trials (1991-present), CINAHL Plus with Full Text (1937-present), SportDiscus with Full Text (1930-present) and Web of Science (1945-present). The last research was performed on December 22, 2021. The following keywords were used to search the concept of physical activity: “aerobic training” OR exercise* OR “physical activity” OR “physical training” OR “resistance training” OR sport* OR “strength training” OR “weight training” OR weightlifting OR “weight lifting” OR running OR football OR hockey OR athletes OR exertion OR “athletic performance”. For olfaction, the following keywords were used: odor OR odour OR aroma OR fragrance* OR smelling OR smell OR olfactory OR olfaction OR normosmic OR normosmia OR hyposmic OR hyposmia OR anosmic OR anosmia OR odorant* OR sniff OR sniffing.

Included studies had to meet the following inclusion criteria: (1) investigate the effect of PA on olfactory acuity; (2) involving humans, both adults and children. On the other hand, studies were not included if (1) olfactory acuity had changed following head trauma or surgery; (2) studies were not completed or accepted in peer-review journals; (3) studies were written in a language other than English or French.

### 2.2 Study Selection

Figure 1 shows the evolution of the number of articles retained and excluded at each stage. Duplicates were removed after studies identification. The first screening of articles was done only using titles by two authors (MC, AD). The second screening of articles was done using abstracts by two authors (MC, AD). Subsequently, a final sorting was done by two authors (MC, AD) using the full texts of the remaining studies. All authors’ conflicts were discussed internally and resolved by a third author (ACG).

**Figure 1.**
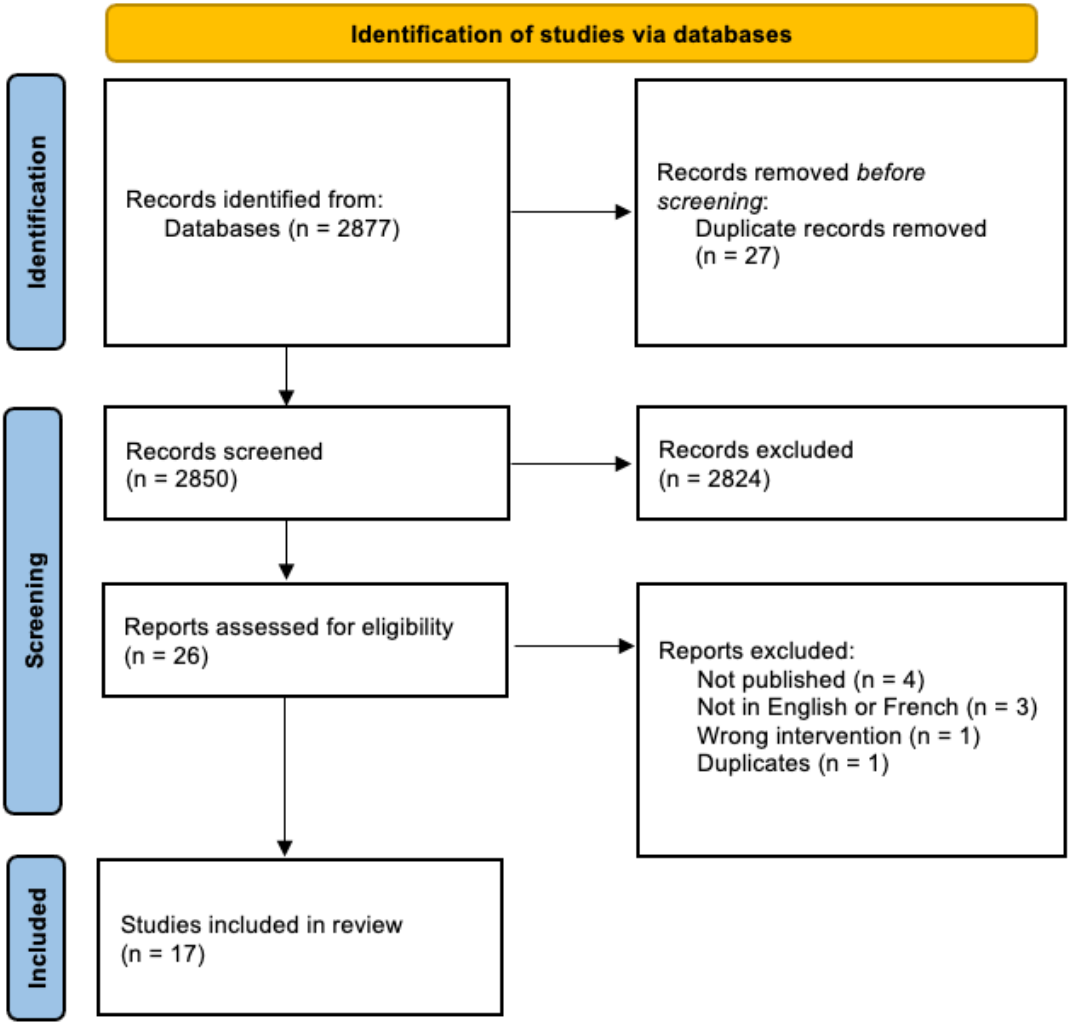
Flowchart of studies selection from their identification to final inclusion

### 2.3 Data Extraction and Analyses

Articles included following the selection process were read and analyzed by two authors (MC, AD). The data were processed by MC, extracted by MC and validated independently by AD. If differences were present, the authors reached a consensus following a discussion. If necessary, a third author (ACG) was consulted to make the decision. Data from included studies are summarized in Table 1, and their protocols are summarized in Table 2.

**Table 1.**
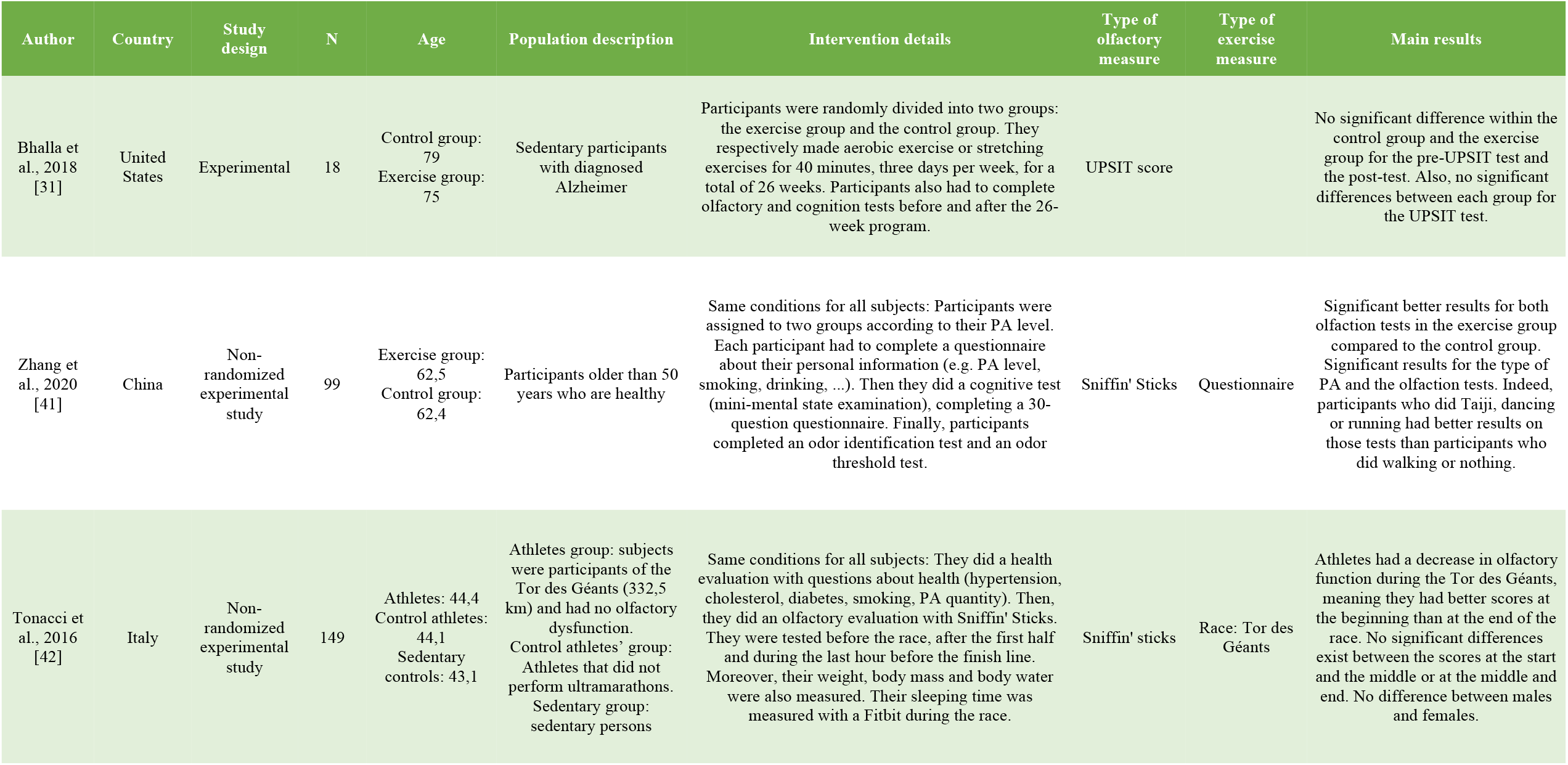
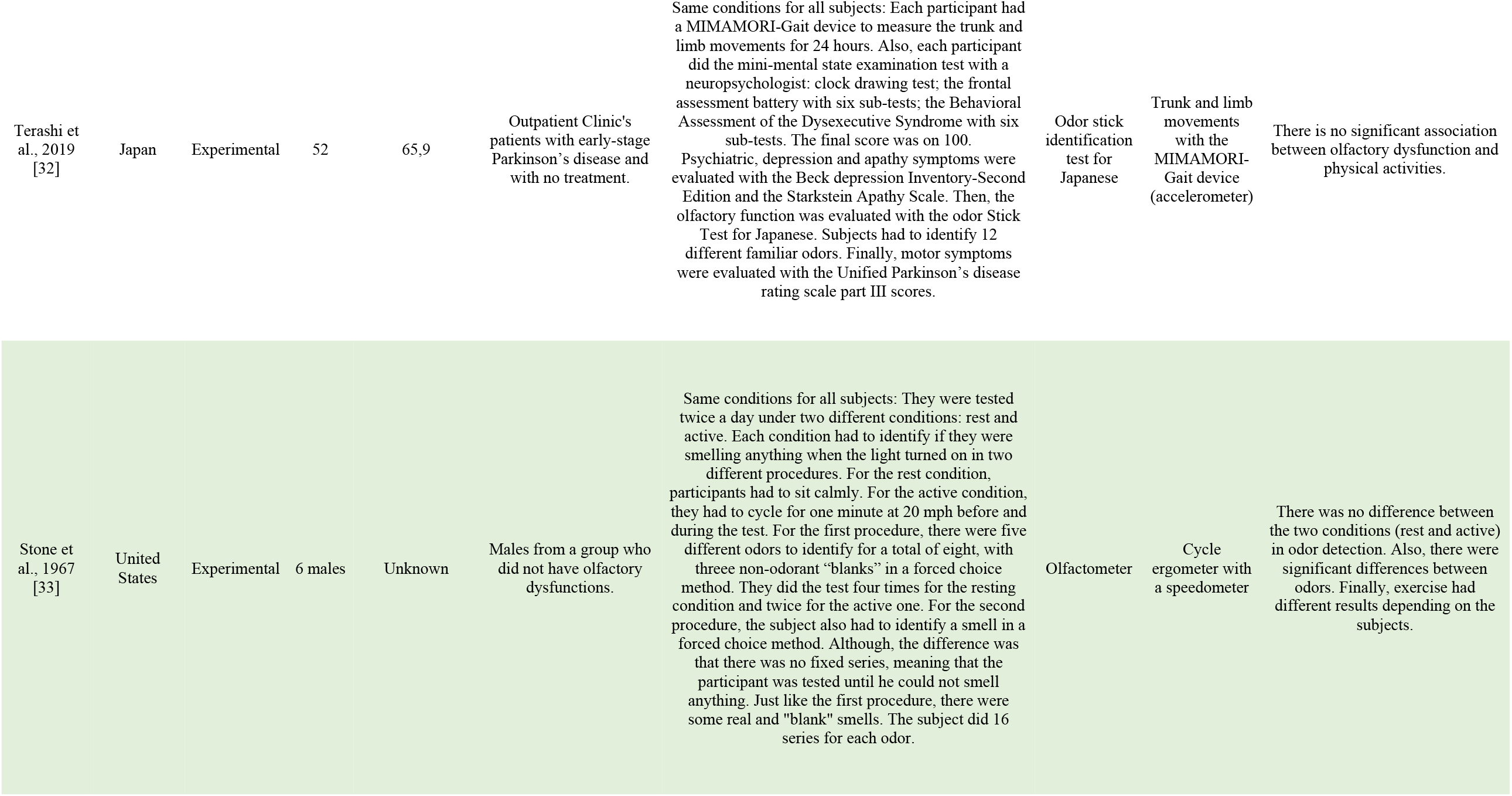
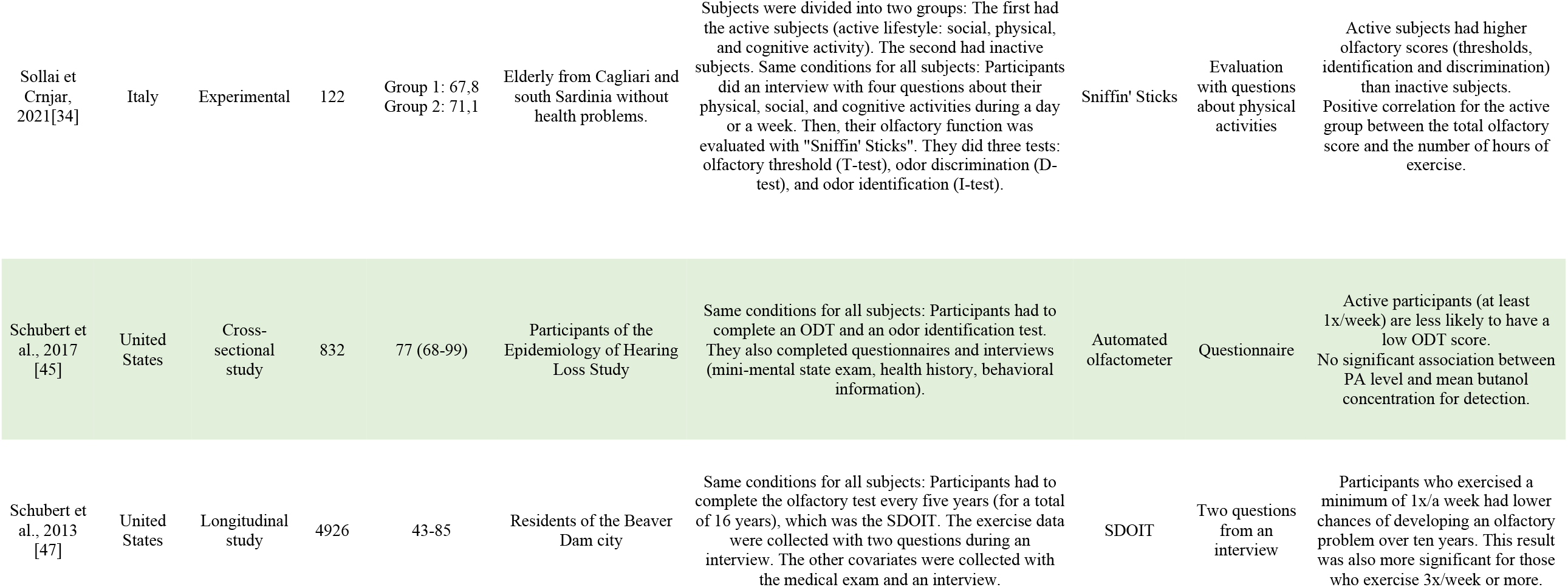
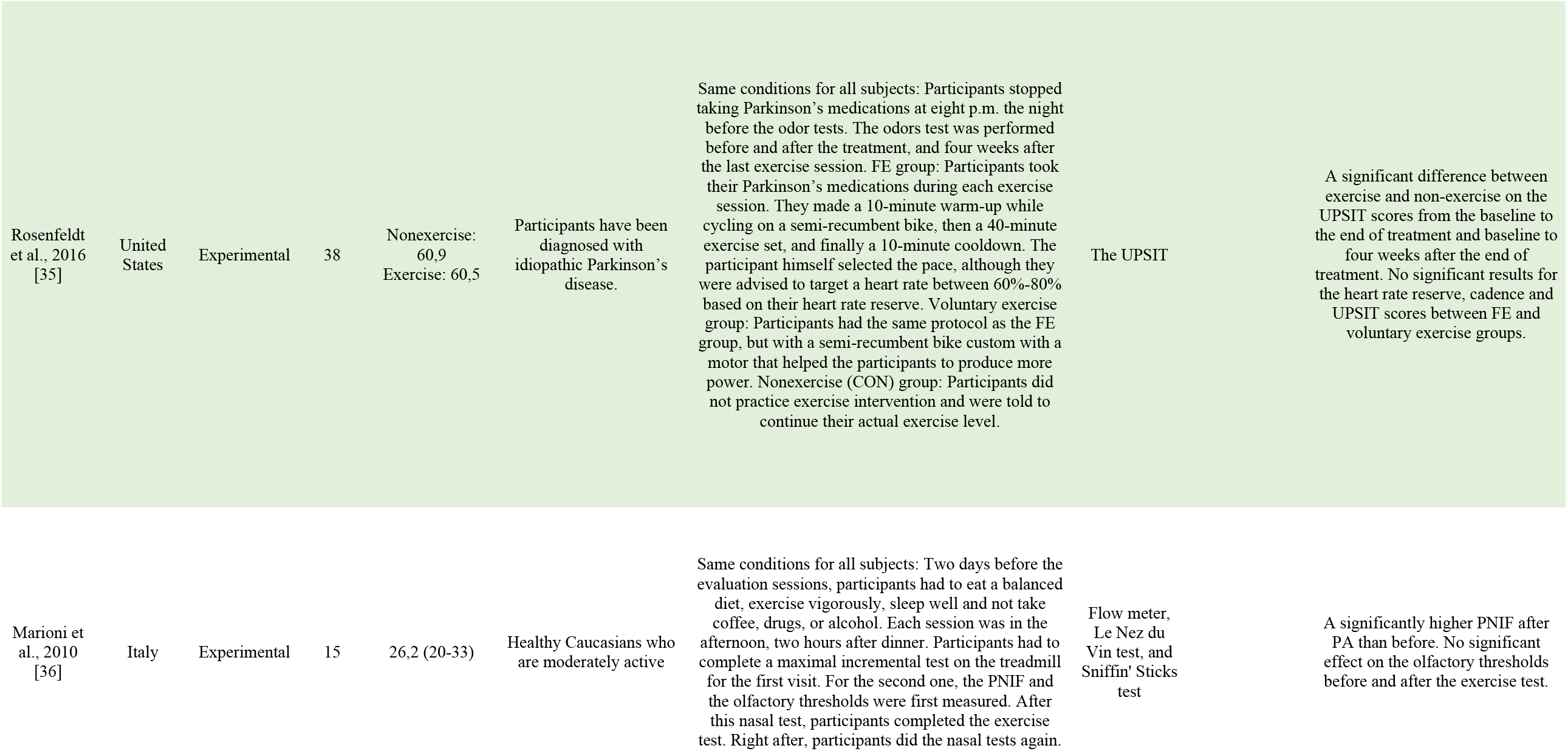
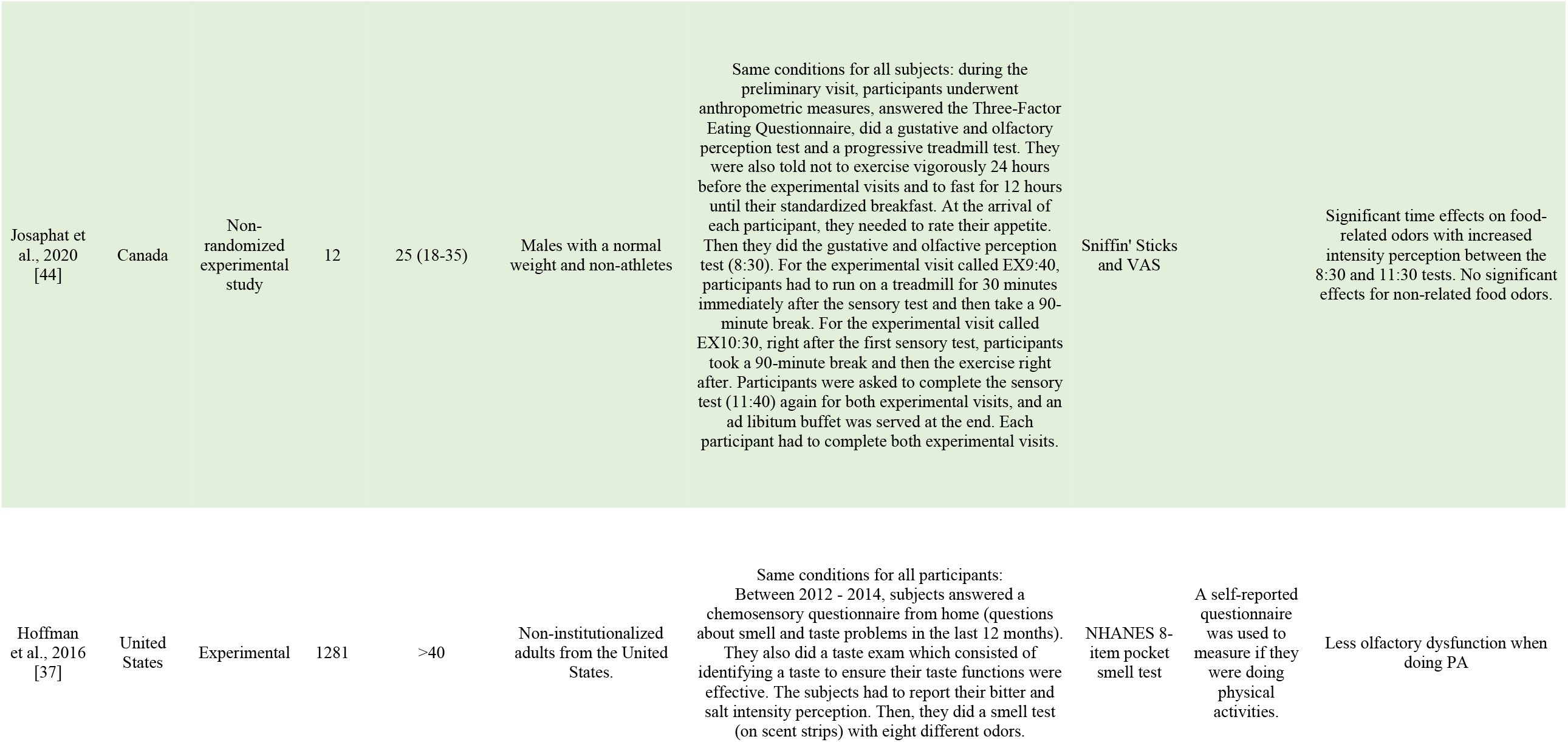
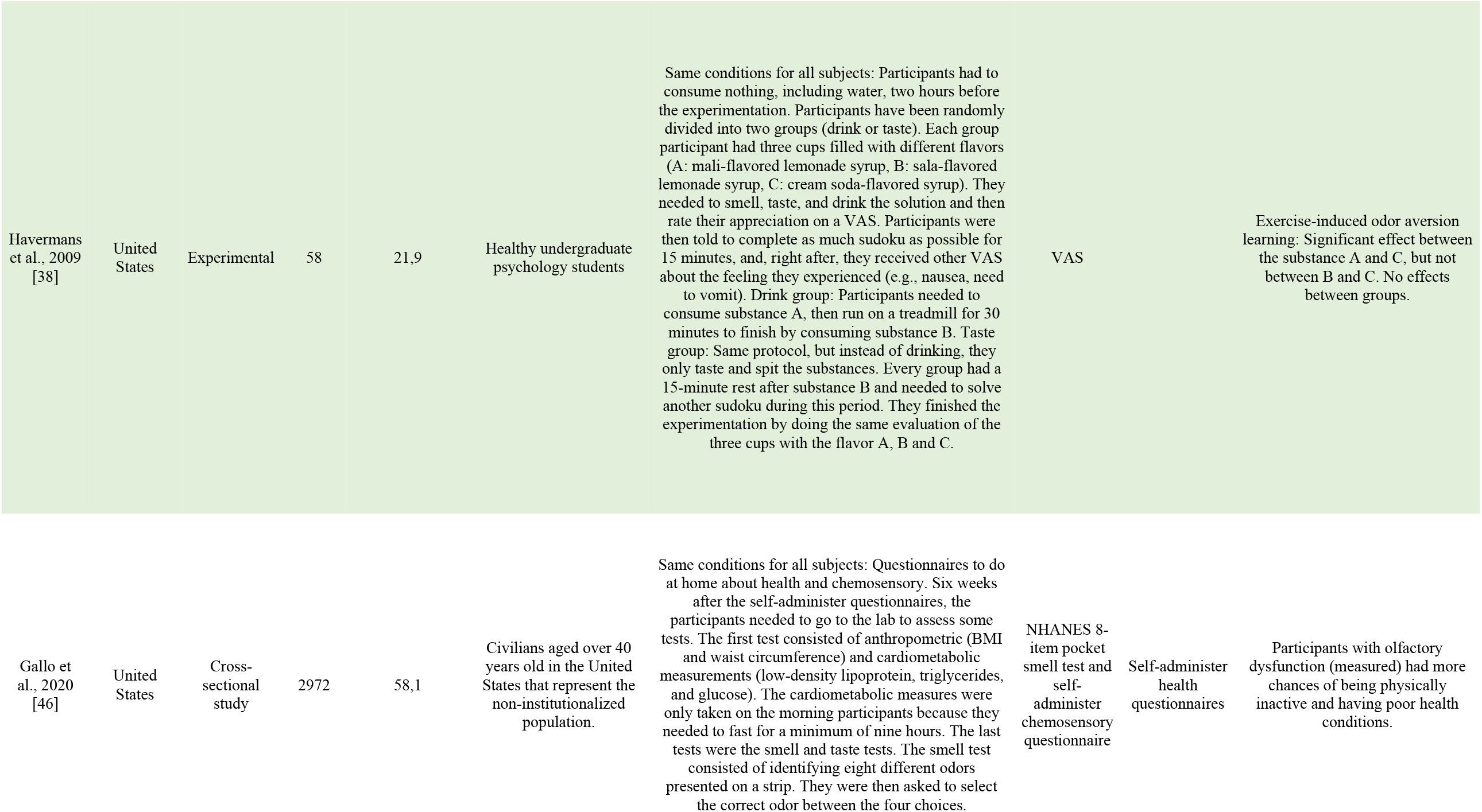
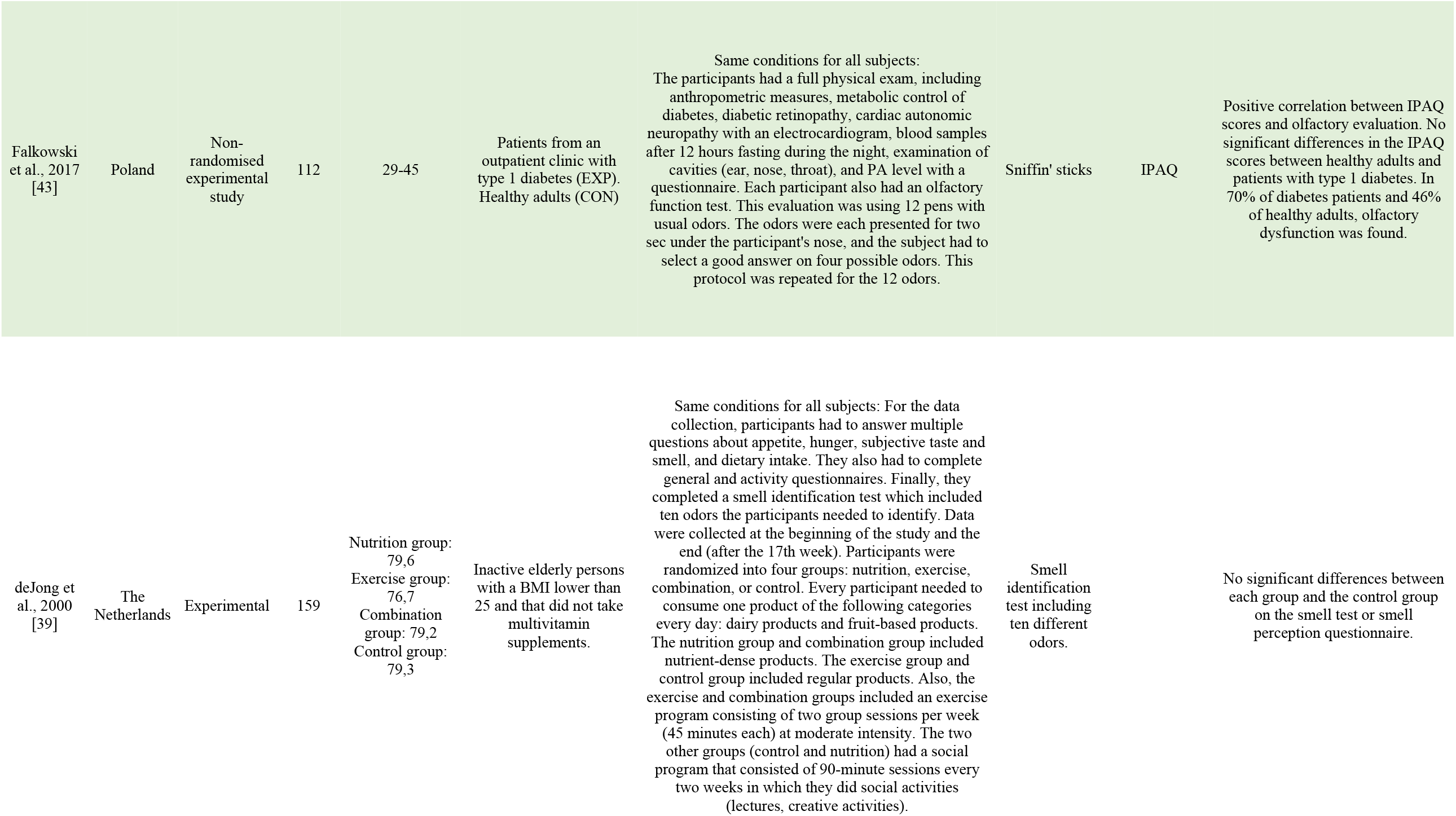
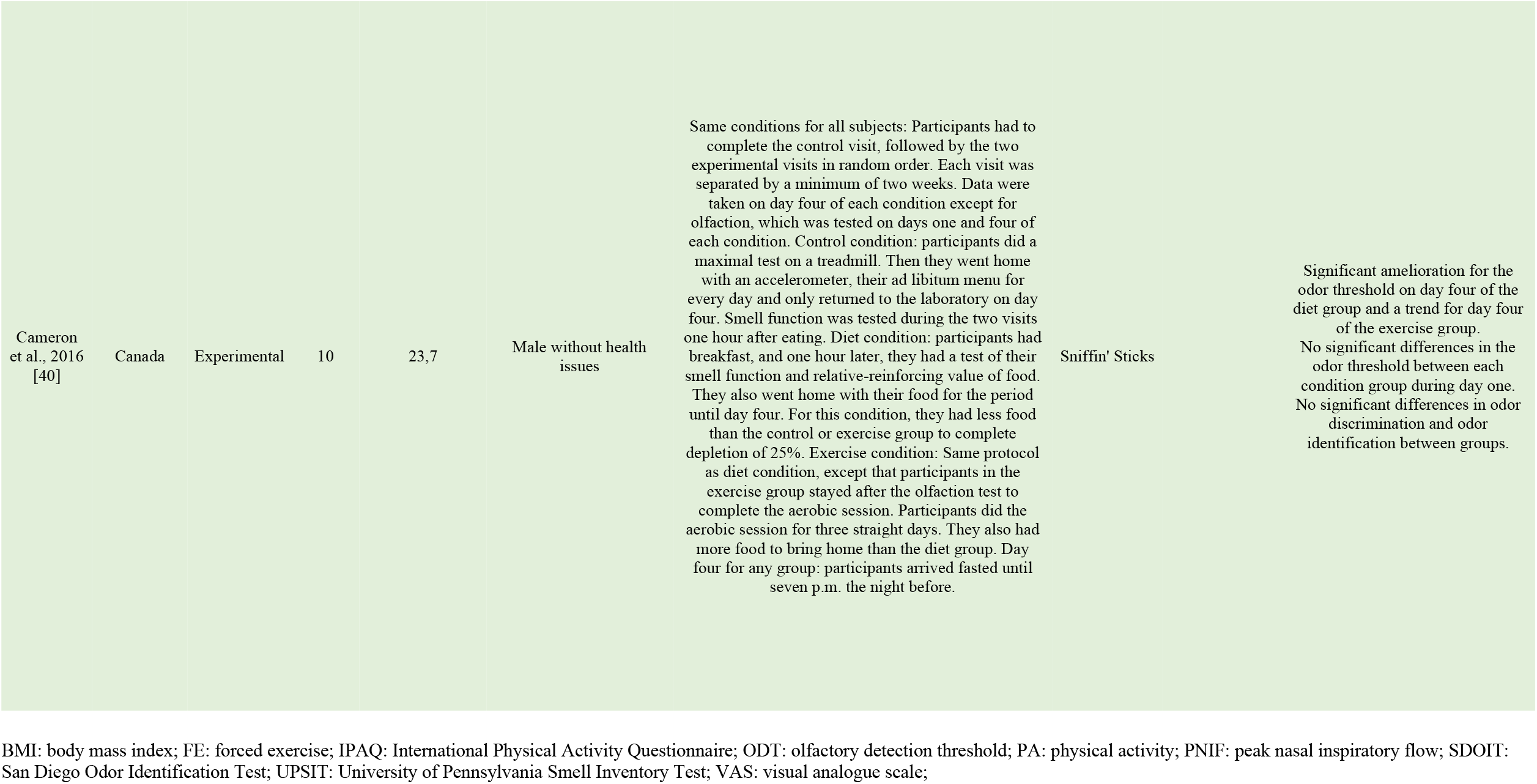
Included studies’ characteristics

**Table 2.** Protocol description

| Author | Variables measured | Type of exercise description | Olfactory experimental protocol | Type of olfactory tests description | Tests duration |
| --- | --- | --- | --- | --- | --- |
| Bhalla et al., 2018 [31] | Olfaction, and cognition | Exercise group modality: On three non-consecutive days per week, participants had to do a 20-minute walk on a treadmill at 1.7 mph with speed increasing to reach 40-55% of their maximal heart rate during the first four weeks. Speed and duration increased every week to reach new intensities and to reach a 40-minute exercise. For example, for weeks 5-18, participants had to reach 50-65% of their maximal heart rate; for weeks 19-26, they needed to be at 60-75%. Stretching group modality: Participants had to complete nonaerobic exercises such as core strengthening and yoga (different each week). They needed to keep their heart rate under 100 bpm. | Participants had to complete the UPSIT test before and after the 26-week program. This test included 40 different odors, and participants had to inhale the odor stick that was presented to them. | Participants needed to choose the correct answer between the multiple choices. Each good answer gave them one point for a total of 40 points. | 26 weeks |
| Zhang et al., 2020 [41] | Mini-mental state examination score, odor identification score, odor threshold score, and personal information (PA level, sex, age, weight, smoking, drinking, education, drug use, surgical history) | Participants who did 30 minutes of PA more than 3x/week were in the exercise group. | Odor identification test modality: Participants had to inhale 16 odorants well known by China's population. The two nostrils were tested. When participants inhaled the odor presented on the Sniffin' Stick, they needed to choose the correct odor.<br>Odor threshold test modality: Participants had 16 solutions of n-butanol to inhale. Each solution was presented with two other sticks without odor. Participants had to choose the stick with the odor between the three choices. | Odor identification test: Participants had to choose the correct odor. One point was given for each good answer for a total of 16 points.<br>Odor threshold test: Participants had to find the stick with the n-butanol solution. One point was given for each good answer for a total of 16 points. | 1-2 hours |
| Tonacci et al. 2016 [42] | Olfactory function, body composition, and sleep. | Subjects completed the race: Tor des Géants in 150 hours maximum. This race included 332,5 km with a 24000 meters positive elevation. | 16 Sniffin' sticks presented to the athlete before the race, during it and one hour before the finish line. The subjects had to choose the good odor of each 16 different Sniffin sticks between four choices. | After being exposed to the odor, the participant had to choose the good one between four choices. Each good answer gave one point for a total of 16. | 3 sessions |
| Terashi et al., 2019 [32] | Visual, frontal and space function, behavior, depression, olfactory function, and motor symptoms. | Participants had to wear the MIMAMORI-Gait device for 24 hours during their routine. The data collection started at ten a.m. and stopped at ten a.m. the next day. They were asked to continue their normal routine and avoid exceptional physical exercise. | Twelve odor sticks with familiar odors were presented to the subject. | After the exposition to the odor sticks, they had to identify the correct odors. | 1 session |
| Stone et al., 1967<br>[33] | Olfactory function | They had to cycle at 20 mph in free wheel one min prior to the odor test and continue the same exercise during the entire odor test. | The subject was in an air-dilution olfactometer where odors were diffused. Each smell was presented for ten sec. There was also a 15-sec interval between each odor presentation. The five odors were heptanol, octanol, cyclohexanone, methyl isobutyl ketone, and methone. "Blank" smells were also presented. Each session included four estimates of the detection thresholds for each participant. At the end of the four sessions, each participant had 16 estimates of the detection thresholds, eight in each condition. | After exposure to the odor, the subject had to say if he detected or not the odor presented. | 4 sessions |
| Sollai and Crnjar,<br>2021 [34] | Olfactory function, physical, social, and cognitive activities | Their level of PA was evaluated with a succession of four questions. | Test one (olfactory threshold): 16 groups of three pens. Each group had two pens with a solvent and the third one with an n-butanol. The goal was to identify the pen with n-butanol. If the subject identified it two times in a row, a reversal of the scale begins. The end of the test happened when the seventh scale was completed. Test two (odor discrimination): 16 groups of three pens with two with the same odor and one with a different. The goal was to identify the one with the different odor.<br>Test three (odor identification): 16 pens with familiar odors. Every time after smelling a pen, they had to choose one item in a force-choice procedure. The addition of the three tests was the final score. | Test one (olfactory threshold): after exposition to the three pens, the participant had to choose the one with an n-butanol odor. Test two (odor discrimination): after exposition to the three pens, the participant had to identify the one with the different odor. Correct answers (0 to 16). Test three (odor identification): after exposition to each pen, the participant had to choose 1 item in a force-choice procedure. Correct answers (between 0 and 16). For the total, they added up the results of the three tests. | 1 session |
| Schubert et al.,<br>2017 [45] | Height, weight, BMI, ODT, PA level, mental state, and health history |  | ODT test modality: Participants had to inhale different concentrations of butanol solution (level 1 = strongest; level 13 = weakest). They started the test at level 8, and they received two puffs, one without odor and the odor with the solution. They had to choose the puff with the odor. If they had it right two times in a row, they went to level 9, which is a weaker solution. On the other hand, if they had it wrong, they went two levels stronger (e.g., level 6). The test stopped when they had three errors in a row or four consecutive level 1 presentations. Odor identification test modality: Every 45 seconds, participants inhaled a different odor a total of eight times and had to identify what it was. | ODT score was the weakest solution that the participant has been able to detect. It was calculated with the mean of the levels of its last two errors. Odor identification: Participants had to choose the correct answer after inhaling the odour. Each good answer gave them one point for a total of eight points. | Data collection was included in the 21-year follow-up examinations of the Epidemiology of Hearing Loss Study. |
| Schubert et al.,<br>2013 [47] | Height, weight, BMI, hemoglobin A1C, education, lifestyle, medical history, olfactory function, and exercise quantity |  | Participants had to complete the San Diego Odor Identification test, which included eight odors. Each odor was presented with a delay of 45 seconds between them. | They had to choose the correct answer between 20 odors, including the eight common odors tested. If they had the wrong answer, they were given the correct one, and the odor was presented a second time later in the experiment. The score represents the number of good identifications after two trials, for a total of eight points. | 1 visit almost every 5 years for 16 years (Total = 3 visits) |
| Rosenfeldt et al.,<br>2016 [35] | Heart rate reserve, cadence, and olfaction scores | Exercise modality: Cardiopulmonary exercise test on a fixed bike at a self-selected pace, including a 10-minute warm-up, 40 minutes exercise set and a 10-minute cooldown. | Participants took the UPSIT test. This test had 40 "scratch and sniff" odors. This test was performed before and after the treatment, and four weeks after the last session. | After scratching and sniffing, the participants needed to select the correct answer between four choices. Each good answer gave them one point, for a total of 40 points. | 3x/week for 8 weeks. |
| Marioni et al., 2010 [36] | PNIF, and olfactory thresholds | Exercise modality: The exercise test on the treadmill included a one-minute warm-up, followed by ten minutes at their anaerobic threshold speed. Then, the speed was increased every 30 seconds by 10% until exhaustion. | Before and after the exercise test, participants had nasal tests. During these tests, they tested the PNIF and the olfactory thresholds. For the PNIF, participants had to inhale and exhale as hard and fast as possible, only with their noses. Participants completed a quick test called Le Nez du Vin for the olfactory thresholds. They did the Sniffin' sticks test if they had only one error or less. For the Sniffin' sticks test, 16 odors were used. During the test, triplets were presented with two solvents and the third with an odorant. Triplets were presented for 20 seconds. | For the PNIF, two maximal inspirations were kept (the highest ones). For the Sniffin' sticks test, participants could have scored between 1 and 16. | 2 sessions separated by at least a week |
| Josaphat et al., 2020 [44] | Taste perception, smell perception, appetite sensation, and energy intake | During the preliminary visit, a progressive treadmill test was administered. This test started at 8 km/h, and the speed increased by 0.5km/h each minute. For the experimental visits, participants had to run on a treadmill for 30 minutes at 70% of their VO<sub>2</sub>max, calculated at the preliminary visit. | 12 Sniffin' Sticks were used to test olfactory functions between each experimental visit. Sticks were placed randomly at 2.5 centimeters in front of the participants for approximately 30 seconds. Odors were food and non-food odors. | After the 30 seconds inhalation, they needed to rate the intensity and pleasantness of the smell on a VAS. | 3 sessions |
| Hoffman et al., 2016 [37] | Taste intensity, olfactory function, participants' characteristics, and lifestyles. | Self-reported questionnaires. Participants were active if they did 3x/week at least ten minutes of vigorous PA per session. | Scent strips of eight different odors: strawberry, chocolate, onion, grapes, smoke, gas, leather soap. The NHANES employee had to scratch the strip to release the odor. | After inhaling the odor, participants had to identify the good one between four choices. Each good answer gave 1 point for a total of 8. A score of 5 or lower meant an olfactory dysfunction. | 1 session |
| Havermans et al., 2009 [38] | Taste rating, and odor rating | Exercise modality: 30 minutes treadmill run at 80% of their maximal heart rate. | Three cups with different flavors were presented to the participants. The flavors were mali-flavored lemonade syrup, sala-flavored lemonade syrup, and cream soda-flavored syrup. Each of these flavors was added to water for a ratio of 1/10. Participants had to smell each cup, then taste it and drink it. This protocol was done at the beginning and the end of the experimentation. | After the actions for each cup, participants had to complete VAS to rate how much they liked or disliked the odor and the taste. | 1-hour session |
| Gallo et al., 2020 [46] | Olfactory function, BMI, waist circumference, triglycerides, glucose, low-density lipoprotein, demographic characteristics, health conditions, PA level, and smoking | Exercise quantity was evaluated with self-reported questionnaires. They were physically active if they did at least ten minutes of moderate-vigorous PA 3x/week. | Participants were presented with strips with different odors (four food odors, two warning odors and two common odors). The strip was placed in front of the participants to sniff it. | Participants needed to choose a good answer between the four choices that were presented to them for each odor stick. Each good answer gave 1 point for a total of 8 points. | 6 weeks |
| Falkowski et al., 2017 [43] | Olfactory function, IPAQ Scores, BMI, low-density lipoprotein cholesterol, high-density lipoprotein cholesterol, triglycerides, Type 1 diabetes duration, and glycated hemoglobin A1C | PA level was evaluated with the IPAQ. | Twelve odors pens (sniffin' sticks) were presented for two seconds, two centimeters under the participant's nose. Each stick has a well-known odor. | After the exposition of the odor, the participant needed to choose the good odor between the four choices. Each good answer gave 1 point for a total of 12. A final score under 11 meant a significant olfactory dysfunction. | 1 session |
| deJong et al., 2000 [39] | Smell, appetite, taste perception, and feeling of hunger | Exercise quantity: Participants had to complete an activity questionnaire (PASE). Some groups (exercise and combination) also performed PA twice a week in a group for 45 minutes at a moderate intensity. The exercises focused on strength, coordination, flexibility, speed, and endurance. | The smell experimentation included ten different odors. Each odor was presented to the participants in a 40mL plastic jar with holes in the lid. | After inhaling the odor, participants had to identify the odor by choosing the correct answer from the list containing 20 choices (ten tested odors and ten random choices). Each good answer gave 1 point, for a total of 10 points. | 17 weeks |
| Cameron et al., 2016 [40] | Height, weight, body composition, attitude toward eating, VO<sub>2</sub>max, energy expenditure, leptin, ghrelin, energy intake, appetite, palatability, relative-reinforcing value of food, and olfaction | Exercise modality: Participants had to complete the aerobic exercise for three straight days. The exercise was at 50% of their VO<sub>2</sub>max until they reached a 25% depletion. | Participants had three tests to complete: Participants were presented with three sticks. Only one of them had an odor (n-butanol). They needed to identify the odorous one for 16 triplets. The second test also presented three sticks simultaneously to the participants. Although they all had an odor this time, two sticks had the same, and the other had a different odor. They needed to identify the stick with the unique odor for 16 triplets. The last test consisted of smelling one stick and choosing the correct answer between four choices. They repeated this test for 16 sticks. | For each test, participants had 1 point if they had the correct answer and no point if they had the wrong answer. Each test had a total of 16 points. | 3 sessions separated by at least 2 weeks between each session. |

BMI: body mass index; IPAQ: International Physical Activity Questionnaire; ODT: olfactory detection threshold; PA: physical activity; PNIF: peak nasal inspiratory flow; UPSIT: University of Pennsylvania Smell Inventory Test; VAS: visual analogue scale; VO<sub>2</sub>max: maximum oxygen uptake;

### 2.4 Risks of Bias and Quality Assessment

Risks of bias and quality assessment were analysed using the QUADAS-2 tool [30]. This tool consists of four key domains, which cover 1) patient selection, 2) index tests, 3) reference standards, and 4) flow of patients through the study and timing of the index test(s) and reference standard (“flow and timing”) [30]. All domains were assessed in terms of their risk of bias. Patient selection, index tests, and reference standards were also assessed in terms of concerns regarding applicability. The risk of bias was assessed by one author (ACG), and a consensus was reached through discussion with two authors (MC and AM).

## 3 Results

### 3.1 Study Selection

Two thousand eight hundred seventy-seven studies were screened based on their titles, of which 26 were kept for full-text studies assessment. Of those studies, 17 were analyzed, and data were extracted. As presented in Table 1, the latter had between 6 and 4926 participants, which gives a total of n = 10 861 participants. There were also 14 of the 17 selected studies with an experimental design [31-44], including non-randomized studies [41-44]. The three remaining studies had a cross-sectional [45, 46] or a longitudinal [47] design. All 17 studies assessed the effect of PA on odor detection thresholds, odor identification, and/or odor appreciation/intensity for different type of odors.

### 3.2 Study and Physical Activity Intervention Characteristics

As presented in Table 1, studies took place in Canada [40, 44], China [41], Italy [34, 36, 42], Japan [32], the Netherlands [39], Poland [43], and the United States of America [31, 33, 35, 37, 38, 45-47]. The majority were conducted on healthy participants [33, 34, 36-42, 44-47] or participants living with a disease (e.g., Parkinson’s Disease)[31, 32, 35, 43]. Each study had an intervention time ranging from one session [32, 34, 37, 41, 43] to a 26-week program [31]. There was also one study with a 21-year follow-up [45]. Some exercise interventions consisted of cycling on an ergometer or running on a treadmill, and the intensity of the exercise was often based on the participant’s maximal oxygen consumption [31, 33, 35, 36, 38, 40, 44]. Other exercise interventions were the amount of daily movement that was associated to olfactory function [32], and an ultramarathon race (e.g., Tor des géants) [42]. Finally, the remaining studies assessed PA levels with a questionnaire (e.g. International Physical Activity Questionnaire [43]), and therefore included various types of PA [34, 37, 41, 43, 45-47].

### 3.3 Olfactory Protocols and Tests

As presented in Tables 1 and 2, 13 out of the 17 studies used sniffin’ sticks or odor pens with different odors [31, 32, 34-37, 40-44, 46, 47]. Two studies presented the odors in jars [38, 39]. The two remaining studies used a flow olfactometer [33, 45]. Olfactory tests included 8 to 40 odors. The odor detection threshold tests mainly included unfamiliar odors such as n-butanol and heptanol. Participants usually had two or three sticks presented to them, and they had to identify the one with the odor. The studies with a flow olfactometer [33, 45] had a different protocol in which participants were asked if they detected the odor or not. For the odor identification tests, most of the odors presented were familiar ones (e.g., strawberries, smoke). Participants had to identify the odor inhaled between various odors shown on a list. Finally, two studies investigated the appreciation/intensity of odors [38, 44] with visual analogue scales. Participants had to rate those variables following an odor exposure.

### 3.4 Effect of PA on the Three Components of Olfactory Acuity

#### Odor detection threshold

Of the 17 studies, six of them tested odor detection thresholds [33, 34, 36, 40, 41, 45]. While two studies showed no significant effect of PA on odor detection thresholds [33, 36], all the others showed significantly higher scores for the odor detection thresholds for the people who were physically more active (e.g., better detection threshold scores by 15% [41]).

#### Odor identification

Twelve studies explored odor identification [31, 32, 34, 35, 37, 39-43, 46, 47]. Four of these 12 studies showed no significant effect of PA on odor identification [31, 32, 39, 40]. Although one study showed a significant decrease in odor detection after a long PA session, more specifically after an ultramarathon of 332,5 km [42], the seven other studies reported a significant increase in odor identification in the physically active group (e.g., better identification scores by 13.12% [41]) [34, 35, 37, 41, 43, 46, 47].

#### Odor appreciation/intensity

In two studies in which appreciation and intensity of odors were assessed, it was revealed that an acute PA session significantly increased the perceived intensity and appreciation aversion [38, 44]. Nevertheless, these results seem to be odor-dependent, and the effect might change with different odors. For example, PA significantly increases odor intensity with food-related odors such as coffee and orange, which was not the case for non-food-related odors such as leather and rose.

### 3.5 Risk of Bias Assessment

While most of the studies had non-existent bias risks/applicability concerns, some were deemed as ‘’unclear’’ OR at ‘’high risk’’ of bias/applicability concerns (Table 3). Concerning patient selection, out of the 17 studies included in this review, two had an unclear potential risk of bias [45, 47], and one was considered at a higher risk [33]. Also, concerning reference standards, three studies were deemed unclear for their potential risk of bias [33, 42, 43]. Concerning flow and timing, two were classified as unclear [32, 34]. Concerning patient selection, four studies were categorized as unclear for their applicability concerns [33, 42, 45, 47]. Concerning the index test, only one study was classified at a higher risk for applicability concerns [32]. Finally, concerning reference standards, two were deemed unclear applicability concerns [33, 43]. While there were two instances where studies were classified at a higher risk of bias/applicability concerns [32, 33], their methodology and overall framework still made them eligible for data extraction in this systematic review.

**Table 3.** Risk of Bias Assessment [30]

| Author, year | Risk of Bias |  |  |  | Applicability Concerns |  |  |
| --- | --- | --- | --- | --- | --- | --- | --- |
|  | Patient Selection | Index Test | Reference Standard | Flow and Timing | Patient Selection | Index Test | Reference Standard |
| Bhalla et al., 2018 [31] | (+) | (+) | (+) | (+) | (+) | (+) | (+) |
| Zhang et al., 2020 [41] | (?) | (+) | (+) | (+) | (+) | (+) | (+) |
| Tonacci et al., 2016 [42] | (+) | (+) | (?) | (+) | (?) | (+) | (+) |
| Terashi et al., 2019 [32] | (+) | (+) | (+) | (?) | (+) | (-) | (+) |
| Stone et al., 1967 [33] | (-) | (+) | (?) | (+) | (?) | (+) | (?) |
| Sollai et Crnjar, 2021[34] | (+) | (+) | (+) | (?) | (+) | (+) | (+) |
| Schubert et al., 2017 [45] | (?) | (+) | (+) | (+) | (?) | (+) | (+) |
| Schubert et al., 2013 [47] | (?) | (+) | (+) | (+) | (?) | (+) | (+) |
| Rosenfeldt et al., 2016 [35] | (+) | (+) | (+) | (+) | (+) | (+) | (+) |
| Marioni et al., 2010 [36] | (+) | (+) | (+) | (+) | (+) | (+) | (+) |
| Josaphat et al., 2020 [44] | (+) | (+) | (+) | (+) | (+) | (+) | (+) |
| Hoffman et al., 2016 [37] | (+) | (+) | (+) | (+) | (+) | (+) | (+) |
| Havermans et al., 2009 [38] | (+) | (+) | (+) | (+) | (+) | (+) | (+) |
| Gallo et al., 2020 [46] | (+) | (+) | (+) | (+) | (+) | (+) | (+) |
| Falkowski et al., 2017 [43] | (+) | (+) | (?) | (+) | (+) | (+) | (?) |
| deJong et al., 2000 [39] | (+) | (+) | (+) | (+) | (+) | (+) | (+) |
| Cameron et al., 2016 [40] | (+) | (+) | (+) | (+) | (+) | (+) | (+) |

(+)—Low Risk; (-)—High Risk; (?)—Unclear Risk.

## 4 Discussion

The objective of this systematic review was to document the effect of PA on olfactory acuity. After the selection process, 17 studies were included. For the odor detection thresholds, four studies showed increased scores [34, 40, 41, 45], and two studies showed no significant results [33, 36]. For odor identification, seven showed an increase [34, 35, 37, 41, 43, 46, 47], one study showed a significant decrease following a prolonged PA session [42], and four studies showed no significant results [31, 32, 39, 40]. For odor appreciation/intensity, one of two studies showed an increase in intensity [44], while the other showed a significant appreciation aversion [38].

### 4.1 Odor Detection Thresholds

The effect of PA on odor detection thresholds has been explored by six studies [33, 34, 36, 40, 41, 45]. Half of these studies compared before and after PA intervention [33, 36, 40], while the other half compared an active and inactive group on a daily basis [34, 41, 45]. Four of the six studies revealed that PA increased odor detection thresholds [34, 40, 41, 45]. Studies that compared active and inactive groups on a daily basis all showed significant increases in odor detection threshold in the active group [34, 41, 45]. Also, two of the three studies that compared before and after a single PA intervention in the same group had no significant results [33, 36]. The only study that offered the same comparison with a significant increase had a longer total intervention. Indeed, the total duration of the experimental protocol lasted four days including three straight days of PA sessions, and significant results were obtained during the last day of the testing, which was on the fourth day [40]. All these results suggest that chronic PA would yield a better impact on odor detection thresholds than acute exercise.

Exercise intensity also seems an important variable to consider with the improvement of odor detection thresholds. Indeed, Zhang, Li [41] compared results between different exercise modalities and highlighted better olfactory outcomes for those who practiced dancing (3-11.3 metabolic equivalents (METS); thresholds score: 8.1/16) or running (3.5-23 METS; thresholds score: 7.4/16) in comparison to walking (1.5-3.8 METS; thresholds score: 4.0/16) [48]. Knowing that dancing and running can be performed at high intensity and that high-intensity exercise induces an anorexigenic effect [49, 50], which means a reduced energy intake following exercise, the hormonal changes described thereafter might play a role in this increased odor detection thresholds [3, 51].

Also, with the knowledge that chronic PA can generate weight loss by increasing energy expenditure and modulating some hormones by lowering leptin levels [52, 53], those two factors might play a crucial role in the odor detection threshold’s function. Indeed, in a systematic review by Peng, Coutts [3], there was a negative association between odor detection threshold and participant’s weight. Also, obesity is often associated with metabolic and hormonal dysfunctions, which can negatively impact the olfactory function. More precisely, obesity is positively correlated with lower ghrelin levels [3, 54], which stimulates the olfactory bulb, and with higher leptin, which is an olfactory inhibitor [3]. The differences between studies with a PA intervention (acute) and studies with a physically active group on a daily basis (chronic) might be due to the differences that chronic *vs* acute PA have on weight and hormone levels.

### 4.2 Odor Identification

The effect of PA on odor identification was the most documented olfactory component. In this review, 12 of the 17 studies covered this topic. Five studies compared odor identification before and after a PA intervention [31, 35, 39, 40, 42]. Three of them showed no significant effect of PA [31, 39, 40], and one showed a decrease in odor identification after PA [42]. This negative change in olfaction following an exercise session might have been produced by the extreme effort in this study as participants ran an ultra-marathon (332,5 km). However, this result is not consistent with hormonal changes that happened during and following such efforts. In fact, Roupas, Mamali [55] demonstrated that hormones such as leptin and insulin are significantly lowered following an ultra-marathon. As previously discussed, those changes should increase olfactory sensitivity [3, 51].

From the seven other studies comparing active and inactive groups on a daily basis [32, 34, 37, 41, 43, 46, 47], only one had no significant results [32]. Although acute PA seems to have little to no impact on odor identification, chronic PA produced much more conclusive and positive results. Once again, this effect might be related to weight loss and hormonal changes associated with the regular practice of PA. However, Peng, Coutts [3] identified no changes between different weight groups for odor identification in their systematic review, even though they admitted that the literature included conflicting results. For the hormonal changes, studies demonstrated that higher fasting blood glucose levels and hypertension might increase the risk of olfactory impairment by diminishing ghrelin and increasing leptin levels [3, 28]. Long-term PA might help regulate these hormones and protect the sense of smell [53].

Furthermore, Serby [56] demonstrated that odor identification decreased in patients with Parkinson’s and Alzheimer’s disease compared to healthy people of the same age. Jalali, Roudbary [2] also explained that odor identification varied between Parkinson’s patients and their stages in the disease. In the present review, three studies had patients with neurodegenerative disease, two with Parkinson’s [32, 35] and one with Alzheimer’s [31]. Rosenfeldt, Dey [35] documented a significant improvement of the odor identification score following an aerobic exercise with Parkinson’s patients (e.g., UPSIT scores four weeks after the treatment: the exercise group had +0.2 points and the control group had - 2.7 points versus the baseline evaluation). Alternatively, Terashi, Taguchi [32] showed no association between olfactory dysfunction and physical activity. In these three studies, diagnosed populations were compared altogether. Even though there are only a few studies with neurodegenerative participants, those results are less optimistic than studies with healthy populations that had better odor identification scores after PA. Those non-significant results with neurodegenerative patients suggest that the effect of PA is population-specific. This difference in the odor identification ability between healthy and neurodegenerative patients might be explained by various factors dependent of the specific disease [57]. For Alzheimer’s and Parkinson’s patients, one of the factors might be the higher reduction of the olfactory bulb volume [57-62]. A study by Brown, Cooper-Kuhn [63] showed that PA could not protect against the neurogenesis of the olfactory bulb. This fact could explain the differences observed in term of odor identification in the studies included in this review.

### 4.3 Odor Appreciation/Intensity

With the lack of articles in the literature regarding the effect of PA on odor appreciation and intensity, this review only identified two studies on this topic [38, 44]. These studies compared the results of before and after an intervention, showing a significant increase in odor intensity [44] and odor appreciation aversion [38]. These two studies focused on young participants and used visual analogue scales to measure odor appreciation/intensity, which might explain their consistent results. They also documented significant effect of PA on odor appreciation/intensity with some odors such as coffee and orange and no effect with other odors such as rose, which suggest that the effect of PA intervention is odor-specific. In fact, PA intervention changed the perception of food-related odors only. A study by Sorokowska, Schoen [64] highlighted that the right insula, anterior cingulate cortex, and putamen brain regions involved in odor processing are more active in response to food-related odors than other type of odor, which may explain these results. Indeed, those brain regions play a key role in reward loops, and hormonal changes such as higher ghrelin, which normally happens before eating, can affect those brain regions [64, 65]. Also, as mentioned earlier in the discussion, ghrelin is a hormone which stimulates the olfactory bulb [3]. Therefore, a higher concentration of this hormone could generate odor aversion [66]. However, it is important to note that it is unlikely that a higher ghrelin level is caused by the PA intervention since a systematic review showed that the ghrelin level will change after chronic physical activity practice and not acute [67]. In light of that information, food odor may have the ability to increase the activation of some brain regions while non-food odor would not. Therefore, these results align with the findings of both studies included in this systematic review.

Another potential cause for the effect of PA on odor intensity might be related to the timing of the testing sequence of the olfactory tests. Indeed, as evidenced by Josaphat, Drapeau [44], there was an increase in odor intensity between 8:30 a.m., and 11:30 a.m., which evidences a time-related effect and not a treatment effect which is the PA intervention. Therefore, it seems possible that the delay between the last energy intake and the odor presentation could impact the perception of odor intensity. In the same direction, ghrelin level, an olfactory stimulator [3], increases as the last caloric ingestion is far away [68]. However, more studies on odor appreciation and intensity are needed to explore these hypotheses, and protocols should identify specific timing for PA intervention and odor tests.

### 4.4 How Can We Measure Olfactory Acuity with a Novel Scope and How Can We Produce More Robust Results?

Over the years, studies that investigated the effect of PA on olfactory acuity have used different experimental paradigms. Some standardized the conditions before the experimentation [31, 33, 35, 36, 38-40, 44] while others tested in natural conditions the participant [32, 34, 37, 41-43, 45-47]. Even though the latter has been useful in determining the effect of acute and chronic PA on olfactory acuity, better standardization of the pre-experimentation conditions would generate more robust results. For example, knowing that hormonal changes can affect olfactory acuity [3], the exact time of the day the experimentation takes place should be controlled. This concept has been well documented in the study of Josaphat, Drapeau [44], which evidenced a time effect on the perceived intensity of food-related odors. Also, all participants should have the same relative energy intake before/during the experimentation to limit hormonal variations since olfaction can vary depending on the satiety state [69]. Furthermore, studies should measure participants’ bodyweight and use this bodyweight status for further analysis since hormone levels, such as ghrelin, change with this variable [3, 53].

In addition, future research should use more precise and easily regulated tools in odor testing. Indeed, most studies included in this review used Sniffin’ Sticks and questionnaires. Although these methods are commonly used, perhaps due to their simplicity, a more objective approach could strengthen the results. In this sense, some recent studies have started using an automatic olfactometer and an electroencephalogram to measure brain responses to odor stimuli [70, 71]. Therefore, this method makes it possible to understand better the brain mechanisms linked to the sense of smell while obtaining reproducible results. The same aspect about regulated tools should apply to PA. Indeed, it is also necessary to better monitor exercise to understand better which exercise modality (frequency, intensity, time, and type) is optimal for improving olfactory acuity.

### 4.5 Strengths and Limitations

Most of the studies included in this systematic review were relatively recent with the majority published in the last ∼10 years. This observation made it possible to have consistency in data collection and methods. In fact, every study that assessed the acute effect of PA used aerobic exercise such as running on a treadmill or pedalling on an ergometer, limiting findings to cardiovascular exercises. This review also included two types of PA, chronic and acute, which made possible to cover a larger portion of the literature to highlight the effects of each one. For odor appreciation and intensity, although a very small number of studies explored this concept, they both used a visual analogue scale, considered the gold standard for subjective sensory data collection [72]. The main limitation would be the time of the day at which data collection was made in every study. There was no consistency between each study for this parameter, which may have modulated the participants’ hunger level/hormones during the olfactory stimulations. Also, the measurement of PA could have been more precise since most of the studies used self-reported questionnaires [34, 37, 41, 43, 45-47]. Indeed, a study demonstrated that a questionnaire such as the International Physical Activity Questionnaire-Long Form overestimates PA time and underestimates sedentary behavior time [73]. All those inconsistencies in the methods made the possibility of performing a meta-analysis impossible. Better measurements of PA could help find the impact of its effect on olfactory thresholds, identification, and appreciation/intensity. In addition, studies rarely measured participants’ bodyweight and body composition to compare them with those having a similar bodyweight but different PA levels. Considering that hormone levels can change with PA and with bodyweights, bodyweight status and PA levels should be controlled to provide more robust results [3, 53]. Since this review includes all the studies mentioned in this paragraph, the same strengths and limitations apply to this review.

## 5 Conclusion

Physical activity, especially chronic, can improve olfactory acuity. More precisely, the detection threshold, identification, and intensity of odors are improved with a chronic practice of PA. Concerns about the loss of smell have recently surfaced in the healthcare community. Therefore, studies evaluating the impact of PA on the sense of smell are important to deepen our understanding of this therapeutic intervention on olfactory acuity.

## Data Availability

All data produced in the present work are contained in the manuscript

## Acknoledgement

The authors thank Denis Arvisais for his help with the search strategy.

